# BRCA-DIRECT digital pathway for diagnostic germline genetic testing within a UK breast oncology setting: a randomised, non-inferiority trial

**DOI:** 10.1101/2024.05.03.24306669

**Authors:** B. Torr, C. Jones, G. Kavanaugh, M. Hamill, S. Allen, S. Choi, A. Garrett, M. Valganon-Petrizan, S. MacMahon, L. Yuan, R. Way, H. Harder, R. Gold, A. Taylor, R. Gabe, A. Lucassen, R. Manchanda, L. Fallowfield, V. Jenkins, A. Gandhi, D.G. Evans, A. George, M. Hubank, Z. Kemp, S. Bremner, C. Turnbull

**Affiliations:** Institute of Cancer Research, Division of Genetics and Epidemiology, Sutton, UK; Brighton and Sussex Clinical Trials Unit, Brighton and Sussex Medical School, Brighton, UK; Department of Clinical Genetics, St Georges Hospital NHS Trust, London, UK; Clinical Genomics Department, Centre for Molecular Pathology, NIHR Cancer Biomedical Research Centre, Royal Marsden NHS Foundation Trust, Sutton, UK; Sussex Health Outcomes Research and Education in Cancer (SHORE-C), Brighton and Sussex Medical School, Brighton, UK; BRCA Journey, Patient Representative, Leeds, UK; Clinical Genetics, East Anglian Medical Genetics Service, Cambridge, UK; Wolfson Institute of Population Health, Queen Mary’s University of London, London, UK; Nuffield Department of Medicine, University of Oxford, Oxford, UK; Department of Gynaecological Oncology, Barts Health NHS Trust, London, UK; Department of Health Services Research, Faculty of Public Health & Policy, London School of Hygiene and Tropical Medicine, London, UK; School of Cancer Sciences, Faculty of Biology, Medicine and Health, University of Manchester, Manchester Academic Health Science Centre, Manchester, UK; Prevent Breast Cancer Centre, Wythenshawe Hospital Manchester Universities Foundation Trust, Manchester, UK; Nightingale and Genesis Breast Cancer Centre, Manchester University Hospitals NHS Foundation Trust, Manchester, UK; Division of Evolution, Infection, and Genomic Sciences, The University of Manchester, Manchester, UK; Cancer Genetics Unit, Royal Marsden NHS Foundation Trust, London, UK; Breast Oncology Unit, Royal Marsden NHS Foundation Trust, London, UK

## Abstract

**BACKGROUND:** Genetic testing to identify germline high-risk pathogenic variants in breast cancer susceptibility genes is an important step in the breast cancer diagnostic pathway. To expand capacity and reduce turnaround time, testing is increasingly offered within ‘mainstream’ oncology services, rather than via referral to clinical genetics. However, mainstream capacity is also stretched, as testing is offered to greater proportions of patients. Novel patient-centred pathways may offer opportunity for improved access.

**PATIENTS AND METHODS:** We recruited 1,140 women with unselected breast cancer to undergo germline genetic testing through the BRCA-DIRECT digital pathway; compromising at-home saliva sampling and consenting, with access to a digital dashboard to complete tasks and a genetic counselling telephone hotline.

Ahead of consenting to the test, participants were randomised to receive information about genetic testing digitally (569/1140, 49.9%) or via a pre-test genetic counselling consultation (571/1140, 50.1%). The primary outcome was uptake of genetic testing. We also measured patient knowledge, anxiety, and satisfaction, and conducted a healthcare professional survey.

**RESULTS:** 1,001 (87.8%) participants progressed to receive their pre-test information and consented to testing. Uptake was higher within participants randomised to receive digital information compared with those randomised to a pre-test genetic counselling consultation (90.8% (95% CI: 88.5% to 93.1%) vs 84.7% (95% CI: 81.8% to 87.6%), p=0.002, adjusted for participant age and site). Non-inferiority was observed in relation to all other outcomes evaluated. Usage of the telephone hotline was modest (<20% of participants; 1,441 total minutes, 344 clinical minutes recorded) and, of 37 healthcare professionals surveyed, there was majority agreement that all elements of the pathway were equivalent to current standard-of-care.

**CONCLUSION:** Findings demonstrate that standardised, digital information offers a non-inferior alternative to conventional genetic counselling consultation, and that an end-to-end patient-centred, digital pathway (supported by genetic counselling hotline) could feasibly be implemented into mainstream breast oncology settings.

## INTRODUCTION

Pathogenic variants (PVs) in genes such as *BRCA1*, *BRCA2* and *PALB2* (BRCA-genes) are associated with elevated risk of breast and other cancers (in particular, ovarian cancer), with well-evidenced interventions for early detection and prevention of disease (1–3). Historically, germline genetic testing for the BRCA-genes and other cancer susceptibility genes (CSGs) was limited by requirement for laborious fragment-by-fragment gene analysis, consequent high costs and slow turnaround times, and was typically only initiated following referral to clinical genetics of families with a strong family history of relevant cancers. Next Generation Sequencing has transformed laboratory workflows, thus dramatically improving capacity and turnaround time. However, other elements of the clinical-laboratory pathway remain laborious, meaning complex eligibility criteria remain necessary to restrict the patient volume eligible for testing(4, 5).

Women identified at time of breast cancer (BC) diagnosis as carrying a high-risk PV in a BRCA-gene may elect to reduce their risk by having bilateral mastectomy instead of, or following on from, localised surgery(6). Furthermore, germline BRCA-gene status has emerged as an important therapeutic biomarker informing adjuvant BC therapy (7–9). For example, PARP-inhibitors have recently been approved in the UK by the National Institute of Clinical Excellence (NICE) for HER2-negative advanced (Talazoparib) and early-stage (Olaparib) BC with germline BRCA PVs (10, 11). Consequently, BRCA-gene testing is increasingly offered to BC patients contemporaneous to diagnosis to inform treatment options.

Due to a lack of capacity in clinical genetics and the delay inherent in referral, there has been increasing momentum for delivery for diagnostic genetic testing in the ‘mainstream’ oncology setting(12). Implementation of this has however had variable success and acceptance owing to (i) perceived lack of expertise regarding genetic information-giving by the mainstream clinicians/surgeons themselves, (ii) lack of time in oncology appointments for detailed information-giving, counselling and consenting, (iii) perception of this role being outside the oncology remit, and (iv) navigation of complex eligibility criteria(13).

We hypothesised that most elements of the germline genetic testing process for BC patients were generic and amenable to a more standardised delivery process. We hypothesised that, if required, complex psychological, legal, clinical and/or risk-based information provision and counselling could be provided in a responsive patient-centred model, available throughout the testing process. Such a pathway could minimise the patient-facing time and administrative burden to oncology professionals for delivery of BRCA-testing. We therefore designed a digital pathway termed BRCA-DIRECT, comprising an online digital workflow portal including delivery of information about genetic testing (or ‘pre-test information’) and consent documentation, postal saliva sampling and a genetic counsellor hotline. We conducted the BRCA-DIRECT pathway study in unselected BC patients across breast oncology units in UK hospitals between 2021 and 2023. The study incorporated a randomised non-inferiority comparison of pre-test information delivery via the digital platform versus via a genetic counselling telephone consult.

## METHODS

### Patients

#### Recruitment

Patients were invited to participate in BRCA-DIRECT by clinical or research teams from five breast oncology units within two National Health Service (NHS) trusts in Manchester or London, UK.

A two-stage consent process was required to initiate genetic testing.

Firstly, the patient was provided with information about the BRCA-DIRECT study within clinic. Patients could ‘express interest’ in participating or provide a reason for decline. Those expressing an interest were given a study pack (including research consent form and a saliva sampling kit) to complete at home and return by post.

Secondly, after signing the study consent, participants were invited via email and/or SMS to create an account on the BRCA-DIRECT website, which gave them access to a personalised dashboard, providing an overview of the steps involved in the genetic testing (GT) pathway. The platform automatically notified and enabled completion of time-stamped tasks (see previously published description and figure 1)(14). This included a digital genetic test consent form, only available after receiving the pre-test information (see randomisation).

**Figure 1:**
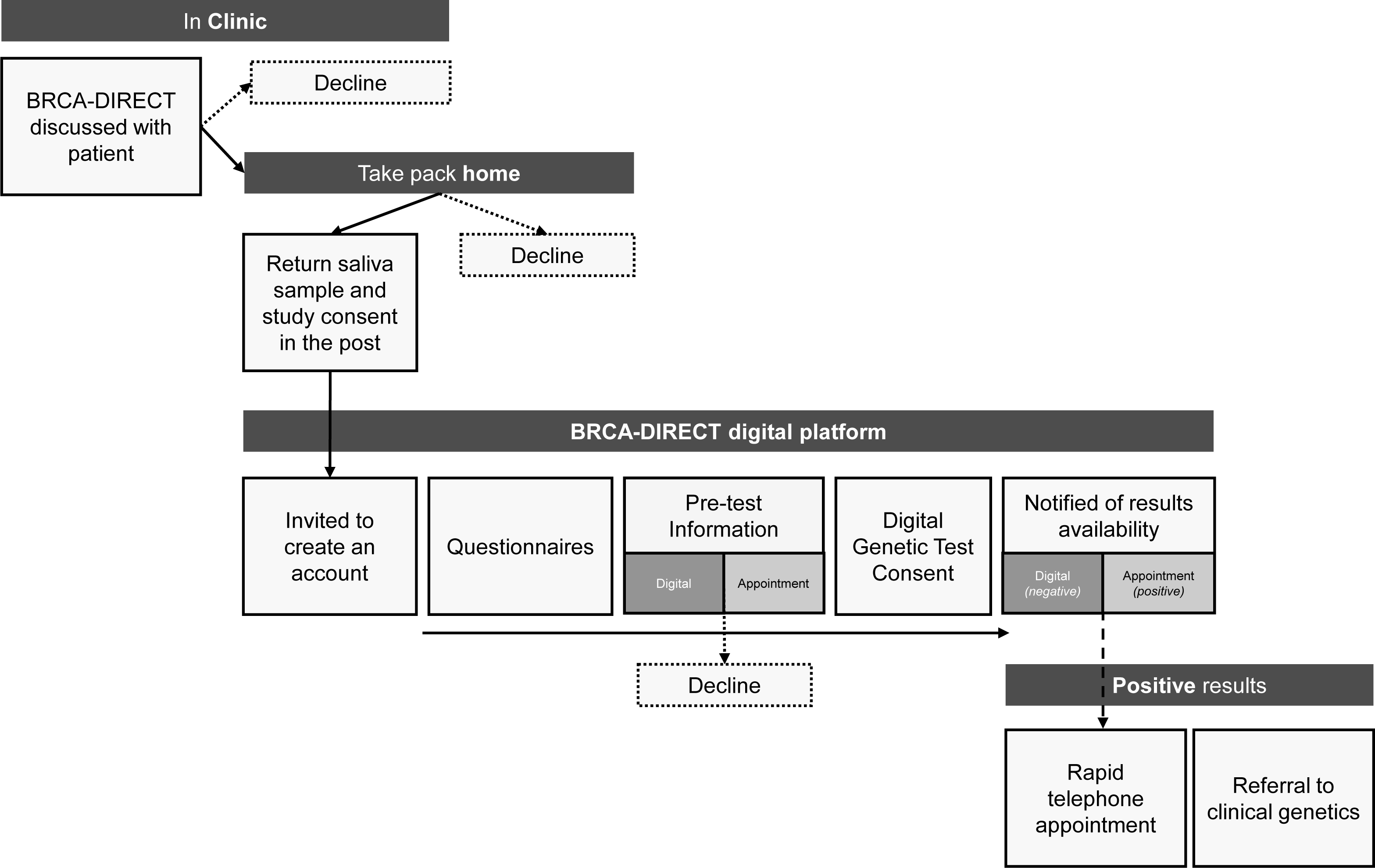
**The BRCA-DIRECT Pathway**

A telephone genetic counselling hotline was available between 9am and 5pm, Monday-to-Friday, through which a genetic counsellor/genetic nurse (GC/GN) could provide support for clinical enquiries, as well as for administrative or technical help.

#### Participant Eligibility

The following criteria were applied:

- Inclusions:

`o` Diagnosis of invasive BC or high-grade ductal carcinoma in-situ (DCIS);
`o` Female;
`o` Over the age of 18 years old;
`o` Good comprehension of English language; and
`o` Access to internet with an email and/or telephone number (this could be via a trusted friend or family-member).
- Exclusion:

`o` Previous BRCA-gene testing.

#### Participant demographics

Participant demographics were collected via a digital survey at baseline. Information relating to BC type and status (newly diagnosed, in follow-up, or metastatic) was collected by local hospital teams at point of consent. Date of primary BC surgery, where available, was recorded prior to study closure.

### Randomisation

#### Pre-test information randomisation

Participants were randomised 1:1 to receive pre-test information digitally via the BRCA-DIRECT website (fully-digital arm) or via a telephone pre-test genetic counselling consultation with a GN/GC (partially-digital arm). Allocation to arm was based on pre-generated, site-specific randomisation lists from the on-line Sealed Envelope randomisation list generator(15). IDs were allocated to participants sequentially as they were registered to BRCA-DIRECT. Participants were only aware of their allocation after they had completed the baseline questionnaires and had proceeded to the point of receiving the pre-test information.

Digital pre-test information consisted of 21 static screens of text and schematics designed to be equivalent in detail and depth to a standard genetic counselling appointment.

Those allocated to receive the pre-test genetic counselling consultation were invited to book an appointment within the next 3-working days (rolling basis). Participants who did not answer the telephone within the slot were notified to rebook an appointment online.

#### Results randomisation

Participants were also pre-allocated to receive results digitally (97.5%) or via a telephone appointment with a GN/GC (2.5%). This randomisation was over-ridden if the participant (i) had a positive genetic test result <u>and</u> (ii) was randomised to receive their result digitally. All participants with positive results, in addition to those randomised to this arm, were issued an online invitation to book a telephone consultation with a GN/GC.

### Study outcomes

#### Non-inferiority of digital pre-test information

The main study aim was to evaluate non-inferiority of digital pre-test information compared with a pre-test genetic counselling consultation.

The primary outcome was uptake of GT. The following secondary outcomes were also evaluated: patient-reported anxiety (State Trait Anxiety Index and Intolerance of Uncertainty); knowledge about genetic testing (14-point study specific questionnaire); and satisfaction with pre-test information delivery (measured on a five-point Likert Scale, within the patient satisfaction survey) (16) (17). More detail on the methods and timepoints for assessment are presented in supplementary table 1, with assumptions used to establish the non-inferiority margins presented in supplementary table 2.

### Feasibility and acceptability outcomes

In addition to non-inferiority outcomes, we aimed to understand broader feasibility and acceptability of the pathway by measuring:

- Overall uptake of the digital pathway, based on expressions of interest and progression to receive pre-test information.
- Uptake of the genetic counselling hotline by consented participants, based on call logs.
- Healthcare professional (HCP) satisfaction with the BRCA-DIRECT pathway. A digital survey was circulated to breast oncology, surgical and clinical genetics HCPs from the recruiting sites between August and September 2022 to understand perception of how elements of the pathway compared to current standard-of-care and views on suitability of the pathway for broader rollout.

We also conducted structured interviews with participants to explore the motivations for, and experiences of, BRCA-testing via the BRCA-DIRECT digital pathway at an early stage in their breast cancer diagnosis and treatment. Findings from these interviews will be reported separately.

### Statistical methods and analyses

#### Sample size

Study sample size (1,000 participants) was calculated to ensure >95% likelihood of identifying at least five individuals with a pathogenic variant at each of the two recruiting trusts, based on a PV-detection rate of 2%. The sample sizes required to achieve 80% power for individual non-inferiority outcomes are presented in supplementary table 1.

#### Analyses

The primary outcome was analysed using a logistic regression model, with fixed effects for randomisation arm and site. Secondary non-inferiority outcomes were analysed using linear mixed effects models with random effect for participant and fixed effects for time point, baseline measure of outcome, randomisation arm and site. Age was added as a covariate to all models a priori to account for the known associations with age and digital accessibility. Baseline outcome measures were included where assessed. Trait anxiety and intolerance of uncertainty scores were included within the model for anxiety.

Sensitivity analyses were performed to identify effects of completing outcome measures outside of time window. Sub-group analyses were performed, where possible, considering the following parameters: localised/advanced BC; genetic test result; reported family history (BC and other cancers <40 years old in first- or second-degree relatives); and method of receiving result.

Analyses were completed following intention-to-treat principles in Stata v17.0.

## RESULTS

### Uptake of digital pathway study for genetic testing

During the recruitment period (05/07/2021 to 15/08/2022 (406 days)), 1,412 patients expressed interest and were provided with a study pack (see CONSORT figure 2). 1,140 of 1,412 (80.7%) patients returned their study consent and saliva sample in the post within the recruitment window, with 569 (49.9%) allocated to the fully-digital arm and 571 (50.1%) allocated to the partially-digital arm. Final follow-up of all participants was completed on 16/01/2023.

**Figure 2:**
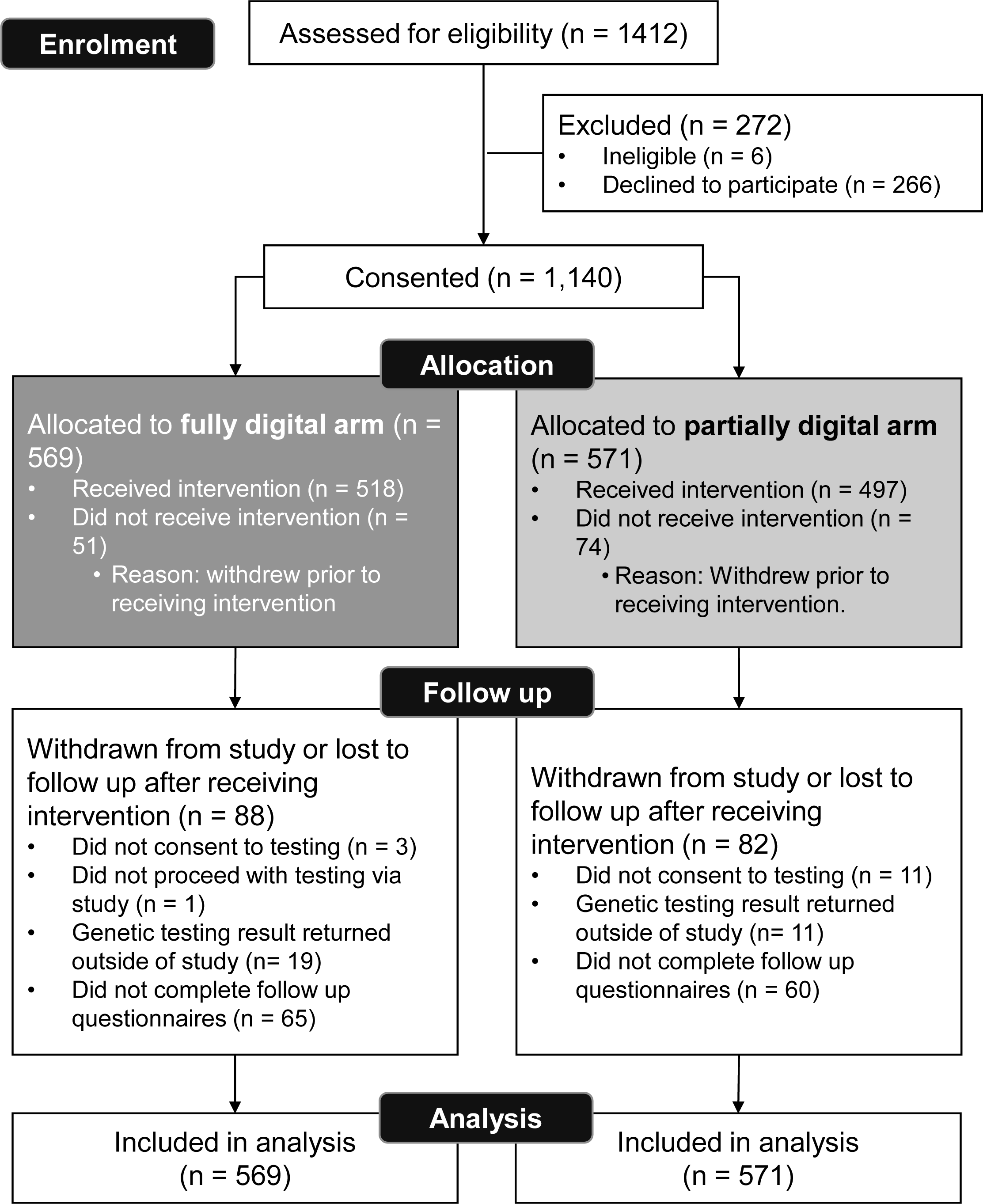
**CONSORT flow chart for recruitment to the BRCA-DIRECT study, including allocation to the fully digital arm (digital pre-test information) or the partially digital arm (genetic counsellor consultation).**

### Participant Characteristics

The mean age (±SD) of participants was 58.6 (± 11.7) years. A majority of participants were white (84.1%), married or partnered (68.4%), and in full or part-time work (57.3%) with 49.9% of participants educated to degree level or higher (see table 1).

**Table 1:** Participant demographics.

|  | Overall (n=1140) |  |  |  |  |  |
| --- | --- | --- | --- | --- | --- | --- |
|  | Partially digital (n=571) |  | Fully digital (n=569) |  | Overall (n=1140) |  |
| | Mean | ±SD | Mean | ±SD | Mean | ±SD |
| **Age** | 58.1 | 11.6 | 59.2 | 11.77 | 58.6 | 11.72 |
|  | n | % | n | % | n | % |
| **Age group** |  |  |  |  |  |  |
| 18-30 years old | 0 | 0 | 1 | 0.2 | 1 | 0.1 |
| 31-40 years old | 31 | 5.4 | 26 | 4.6 | 57 | 5 |
| 41-50 years old | 130 | 22.8 | 113 | 19.9 | 243 | 21.3 |
| 51-60 years old | 177 | 31 | 176 | 30.9 | 353 | 31 |
| 61-70 years old | 146 | 25.6 | 149 | 26.2 | 295 | 25.9 |
| 71-80 years old | 66 | 11.6 | 89 | 15.6 | 155 | 13.6 |
| 81+ years old | 21 | 3.7 | 15 | 2.6 | 36 | 3.2 |
| *Total with available data* | 571 | 100 | 569 | 100 | 1140 | 100 |
| *Missing/prefer not to say* | 0 | 0 | 0 | 0 | 0 | 0 |
| **Ethnicity** |  |  |  |  |  |  |
| White | 418 | 80.5 | 466 | 87.6 | 884 | 84.1 |
| Asian or Asian British | 41 | 7.9 | 26 | 4.9 | 67 | 6.4 |
| Black African/Caribbean or Black British | 20 | 3.9 | 15 | 2.8 | 35 | 3.3 |
| Mixed or multiple ethnicities | 14 | 2.7 | 14 | 2.6 | 28 | 2.7 |
| Other | 26 | 5 | 11 | 2.1 | 37 | 3.5 |
| *Total with available data* | 519 | 90.9 | 532 | 93.5 | 1051 | 92.2 |
| *Missing/prefer not to say* | 52 | 9.1 | 37 | 6.5 | 89 | 7.8 |
| **Highest qualification** |  |  |  |  |  |  |
| no qualifications | 31 | 6.2 | 40 | 7.7 | 71 | 7 |
| GCSE or equivalent | 116 | 23.2 | 105 | 20.3 | 221 | 21.7 |
| NVQ or equivalent | 73 | 14.6 | 60 | 11.6 | 133 | 13.1 |
| A Levels | 35 | 7 | 50 | 9.7 | 85 | 8.3 |
| Degree | 165 | 32.9 | 165 | 31.9 | 330 | 32.4 |
| Higher degree | 81 | 16.2 | 97 | 18.8 | 178 | 17.5 |
| *Total with available data* | 501 | 87.7 | 517 | 90.9 | 1018 | 89.3 |
| *Missing/prefer not to say* | 70 | 12.3 | 52 | 9.1 | 122 | 10.7 |

| Marriage status |  |  |  |  |  |  |
| --- | --- | --- | --- | --- | --- | --- |
| single | 68 | 13.2 | 64 | 12.2 | 132 | 12.7 |
| married or partnered | 349 | 67.9 | 362 | 68.8 | 711 | 68.4 |
| divorced | 60 | 11.7 | 53 | 10.1 | 113 | 10.9 |
| widowed | 37 | 7.2 | 47 | 8.9 | 84 | 8.1 |
| *Total with available data* | 514 | 90 | 526 | 92.4 | 1040 | 91.2 |
| *Missing/prefer not to say* | 57 | 10 | 43 | 7.6 | 100 | 8.8 |
| Employment status |  |  |  |  |  |  |
| unemployed | 34 | 6.8 | 38 | 7.3 | 72 | 7.1 |
| working part time | 111 | 22.3 | 111 | 21.4 | 222 | 21.8 |
| working full time | 188 | 37.8 | 173 | 33.3 | 361 | 35.5 |
| retired | 165 | 33.1 | 197 | 38 | 362 | 35.6 |
| *Total with available data* | 498 | 87.2 | 519 | 91.2 | 1017 | 89.2 |
| *Missing/prefer not to say* | 73 | 12.8 | 50 | 8.8 | 123 | 10.8 |
| Breast cancer status |  |  |  |  |  |  |
| New diagnosis, pre-surgical | 155 | 27.2 | 155 | 27.2 | 310 | 27.2 |
| New diagnosis, pre-surgical, neoadjuvant, chemo | 75 | 13.2 | 59 | 10.4 | 134 | 11.8 |
| New diagnosis, post surgical | 120 | 21.1 | 103 | 18.1 | 223 | 19.6 |
| In follow-up | 188 | 33 | 212 | 37.3 | 400 | 35.1 |
| Metastatic breast cancer | 31 | 5.4 | 40 | 7 | 71 | 6.2 |
| *Total with available data* | 569 | 99.6 | 569 | 100 | 1138 | 99.8 |
| *Missing* | 2 | 0.4 | 0 | 0 | 2 | 0.2 |
| Reported family history of cancer |  |  |  |  |  |  |
| Yes - Family history reported | 232 | 44.6 | 241 | 44.8 | 473 | 44.7 |
| No family history reported | 288 | 55.4 | 297 | 55.2 | 585 | 55.3 |
| *Total with available data* | 520 | 91.1 | 538 | 94.6 | 1058 | 92.8 |
| *Missing* | 51 | 8.9 | 31 | 5.4 | 82 | 7.2 |

### Non-inferiority

#### Consent to and uptake of genetic test

Out of 1,140 participants, 1,001 (87.8%) proceeded to consent to GT (515/569 in the fully-digital arm and 486/571 in the partially -digital arm), whilst 139 (12.2%) did not (54/569 in the fully-digital arm and 85/571 in the partially-digital arm).The adjusted proportions of uptake in each arm were 84.7% (95% CI 81.8% to 87.6%) uptake in the partially-digital arm and 90.8% (88.5% to 93.1%) in the fully-digital arm (p=0.002). The adjusted difference between the arms was +6.1% (2.4% to 9.8%) (supplementary figure 1a, table 2). Therefore, uptake of genetic testing in the fully-digital arm was non-inferior (and also superior) to the partially-digital arm.

**Table 2:** Non-inferiority outcomes.

|  | Observations<br>(participants) | Fully-digital<br>(digital pre-test<br>information),<br>percentage or mean<br>(95% CI) | Partially-digital<br>(pre-test genetic<br>counselling<br>consultation),<br>percentage or mean<br>(95% CI) | Difference<br>(95% CI)<br>P-value | non-inferiority<br>margin and<br>conclusion |
| --- | --- | --- | --- | --- | --- |
| Uptake of<br>Genetic Testing | 1,140<br>(1,140) | 90.8% (88.5% to<br>93.1%) | 84.7% (81.8% to<br>87.6%) | 6.1%<br>(2.4% to 9.8%)<br>$p=0.002$ | 5.5%<br>non-inferior |
| Anxiety | 2,753<br>(994) | 38.3 (37.7 to 39.0) | 37.8 (37.1 to 38.5) | 0.5<br>(-0.4 to 1.4)<br>$p=0.269$ | +3.0<br>non-inferior |
| Knowledge | 1,755<br>(989) | 8.2 (7.9 to 8.4) | 9.2 (9.0 to 9.5) | -1.1<br>(-1.4 to -0.8)<br>$p<0.001$ | -1.4<br>non-inferior |
| Participant<br>satisfaction | 908<br>(908) | 4.7 (4.6 to 4.7) | 4.7 (4.6 to 4.7) | -0.002<br>(-0.1 to 0.1)<br>$p=0.962$ | -0.75<br>non-inferior |

Included within the group who did not consent to GT were participants who failed to register on the platform (51.1%, n=71), did not proceed with the digital baseline activities (38.8%, n=54), or withdrew after receiving the pre-test information (10.1%, n=14).

#### Patient Knowledge about Genetic Testing

Knowledge scores ranged from 0 to 14, representing the proportion of answers scored correctly within the knowledge questionnaire. Knowledge scores increased from baseline in both arms, at both timepoints measured (1-day post genetic test consent and 28-days post results) (supplementary figure 2). Overall adjusted mean knowledge scores were 9.22 (95% CI: 9.00 to 9.45) in the partially-digital arm and 8.15 (95% CI: 7.93 to 8.37) in the fully-digital arm. The adjusted effect of arm on knowledge score was −1.07 (95% CI: −1.39 to -0.75) in the fully-digital arm compared to the partially-digital arm (p<0.001) (supplementary figure 1b, table 2). Based on the non-inferiority (NI) margin (−1.40), the fully-digital arm was non-inferior to the partially-digital arm in relation to knowledge about GT.

#### Patient Anxiety

State anxiety scores, as measured by the State Trait Anxiety Index, ranged from 20 to 80, with a lower score representing less anxiety. Overall adjusted mean anxiety scores were 37.79 (95% CI: 37.13 to 38.46) in the partially-digital arm and 38.31 (95% CI: 37.66 to 38.96) in the fully-digital arm, with anxiety decreasing over time from baseline (supplementary figure 3). The adjusted effect of arm on anxiety score was 0.51 points (95% CI: -0.41 to 1.44) in the fully-digital arm compared to the partially-digital arm (p = 0.277) (supplementary figure 1c, table 2). The NI margin was +3, therefore, the fully-digital arm was non-inferior to the partially-digital arm in terms of reported anxiety.

#### Patient Satisfaction

In both arms, >90% of participants reported satisfaction scores of four or five, out of five, on the Likert scale (1-very unsatisfied; 5 – very satisfied). Adjusted mean satisfaction scores were 4.67 (95% CI: 4.61 to 4.73) in the partially-digital arm and 4.67 (95% CI: 4.60 to 4.73) in the fully-digital arm. The adjusted effect of arm on patient satisfaction score was -0.002 points on Likert scale (95% CI: -0.09 to 0.09) in the fully-digital arm compared to the partially-digital arm (p = 0.962) (supplementary figure 1d, table 2), thus the fully-digital arm was non-inferior to the partially-digital arm in terms of patient satisfaction.

#### Subgroup and sensitivity analyses

There were no effects observed on the outcomes of non-inferiority analyses following sensitivity or subgroup analyses (see supplementary table 3a-d).

### Test-offer-to-results-time

Median (interquartile range (IQR)) turnaround time of results was 54.0 (24.0 to 69.0) days for Manchester and 47.0 (36.0 to 63.5) days for London.

667/1140, or 58.5%, of participants were newly diagnosed with a BC at point of recruitment and of these, 407/667 (or 65.6%) had not yet undergone primary surgical treatment for their BC. 378/407 (92.9%) subsequently proceeded to consent to genetic testing and of these 222/378 (58.7%) received their result before their planned surgery date (where this was known).

### Hotline utilisation

Hotline call logs recorded by the study team covered the period 21/07/2021 to 11/01/2023. During which time, calls were recorded from 201/1140 (17.6%) participants: 90/569 (15.8%) of whom had been allocated to the fully-digital arm and 111/571(19.5%) of whom had been allocated to the partially-digital arm. Overall, 324 hotline call logs were recorded (amounting to 1,441 minutes of calls) of which 50 (15.4%) were clinical and 274 (84.6%) were administrative.

### Healthcare professional satisfaction

Responses were recorded from 37 healthcare professionals (16 Manchester and 21 London). 19/37 (51.4%) respondents were consultant breast oncologists or surgeons with other respondents comprising oncology clinical nurse specialists, research nurses, genetic counsellors, clinical geneticists, and trainee oncologist/surgeons.

On average, healthcare professionals expressed agreement (either 4 (agree to some extent) or 5 (strongly agree)) that all aspects of the pathway were equivalent, or superior, to current standard-of-care, with an overall median score (IQR) of 4.5 (4.3 to 5.0) factoring in all aspects of the pathway (figure 3). The lowest scores were recorded for consideration against current standard-of-care in regard of ‘patient compliance with the pathway’ (4.0 (3.0 to 5.0)) and ‘clinical monitoring of patient progress’ (4.0 (3.5 to 5.0)).

**Figure 3:**
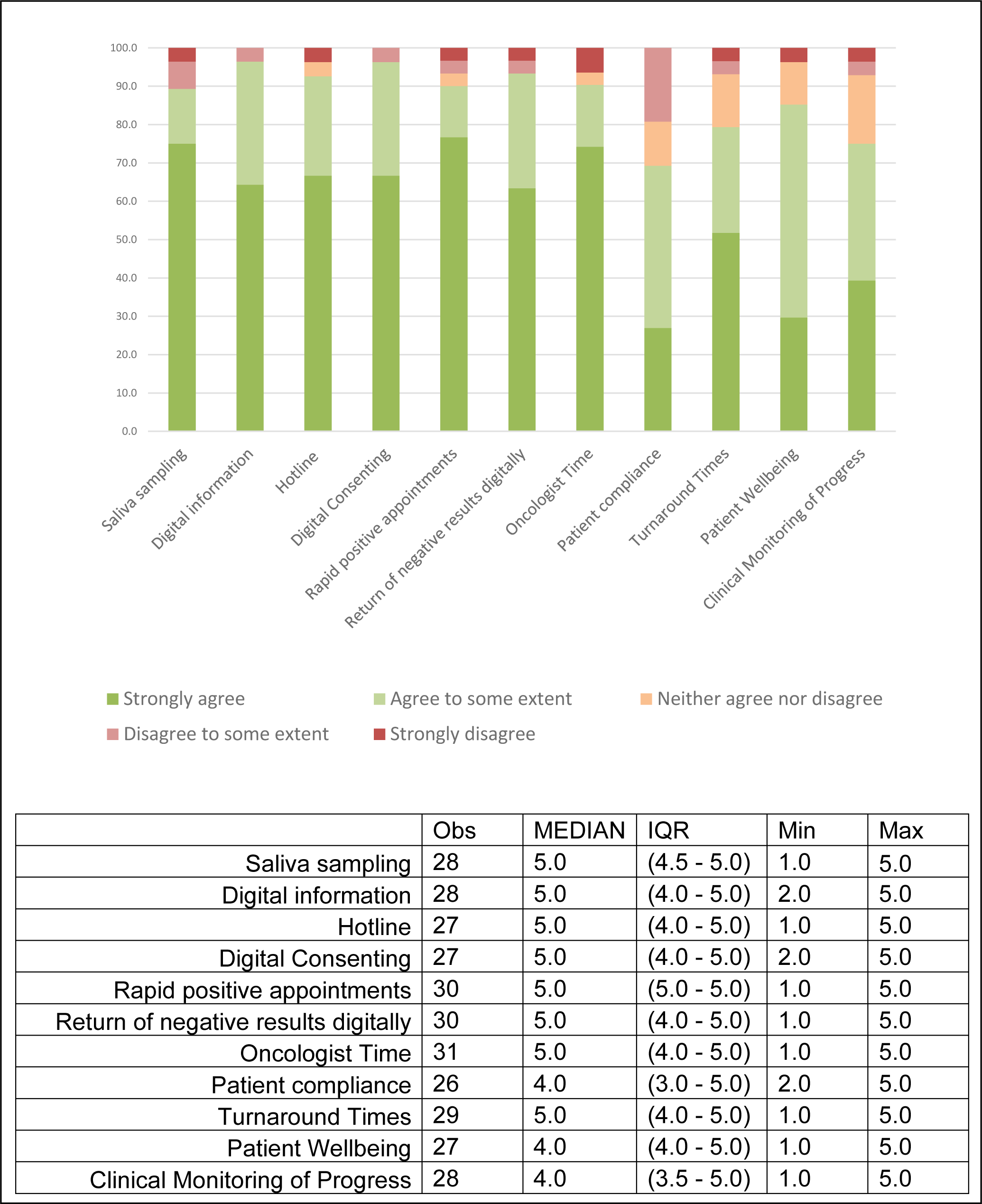
**Healthcare professional reported agreement (1 strongly disagree, 5 – strongly agree), on whether aspects of the BRCA-DIRECT pathway were equivalent (or superior) in comparison to current standard-of-care.**

## DISCUSSION

To our knowledge, this is the largest study reported to date of germline genetic testing using a digital pathway in cancer patients. We have demonstrated logistical feasibility and acceptability of a patient-centred, digital pathway for mainstreaming of BRCA-testing for unselected female BC patients. Via randomised comparison, we also showed non-inferiority against a range of outcomes of digital delivery of pre-test information compared with pre-test genetic counselling consultation.

We observed attrition upstream of participants (i) consenting to the research study and (ii) receiving the pre-test information (19.3% of those initially expressing interest, Figure 2). At both stages, attrition may have partly been a consequence of the demands of the research study through which the BRCA-testing was being delivered. For example, there was need for both research consent as well as standard genetic testing consent, which in some cases proved confusing to patients. Furthermore, being a research study placed additional time requirements on participants for completion of questionnaires. However, this attrition may also reflect barriers to inclusion within the study or progression through the pathway; there was limited opportunity for collection of data within this research study regarding non-participation.

The demographics of participating patients are broadly reflective of BC patients nationally, albeit slightly younger in average age of diagnosis (59 compared with 62 years old). However, there plausibly may be socio-demographic differences in characteristics between all patients presenting in clinic for BC management during the study window and those taking up BRCA-testing. These may reflect biases relating to those offered the study, those expressing interest following offer, and those progressing digitally to the point of test uptake (genetic test consent). Such factors are relevant to the generalisability of the findings; similar factors may well pertain to uptake of BRCA-testing through existing standard pathways and warrant further exploration to improve equity of access to BRCA-testing more generally. However, these biases should not be differential between the two arms of the study, and as such, for those who do opt for testing via a digital pathway, the observation of superior test uptake in the fully-digital arm is of note.

Indeed across all outcomes, our randomised comparison demonstrated non-inferiority of digital delivery of pre-test information compared with a pre-test genetic counselling consultation. Though it is important to consider that our comparator arm was a “partially-digital” pathway, limited to comparison of just the pre-test information, rather than a true standard-of-care arm for BRCA-testing. Nonetheless, our findings relating to equivalence of patient-reported anxiety and satisfaction were promising. Additionally, the relatively low demand for clinical support via the genetic counselling hotline similarly indicate low proportions with substantial anxiety or need for extra support throughout the process.

The genetic counselling hotline was intended to provide optional access to a GN/GC to supplement digital pre-test information. Thus, patient contact with a GN/GC is not necessitated within the proposed pathway but rather allows patient-centred access to support if required. This differs from previous studies in cancer patients, which have largely focussed on implementing digital information or tools in addition (or prior) to genetic counselling (18, 19).

Two exceptions to this, with similar findings, are a US study, reported by Swisher et al., that reported noninferiority of patient distress at three months and no statistically significant differences in anxiety, depression, or decisional regret between participants who did not have mandatory pre-test counselling (received information in the format of a video) compared to those who had pre-test genetic counselling (20). This study was outside of mainstream settings, for participants recruited from the public with a personal or family history of breast or ovarian cancer.

Another study reported by Sie et al. (2013), gave 161 selected BC patients referred to clinical genetics services within the Netherlands the option to proceed with testing via a fully digital route, as alternative to conventional consultation (there was no genetic counselling hotline in this study). Sai et al. also reported high uptake in the digital pathway, that more patients preferred testing without prior face-to-face counselling, and similar outcomes in relation to distress (anxiety) and satisfaction between the digital arm and conventional clinical genetics consultation arm (21).

Within our study, we also compared knowledge between the two arms and our findings reveal that, whilst lower, mean patient knowledge scores in the fully-digital arm lay within the pre-set margin of non-inferiority. Therefore, the standardised material we produced with our patient involvement group can be deemed suitable and sufficient for providing pre-test information generating acceptable knowledge levels, as compared to a GC/GN consult. However, a priority area for future exploration is improving inclusivity and accessibility of digital materials to meet diverse patient needs and to ensure that we are not exacerbating health inequalities. For example, for different learning styles, reading levels, or languages, and accessible content for those with hearing or sight loss, which could be achieved via more interactive tools, use of audio-visual content, or physical paper-based information as an adjunct.

Previous studies of digital pathways have largely focused on community-based ascertainment for genetic testing, for example BRCA founder mutation testing for those with Jewish ancestry. Our study is the first focussed on using a digital pathway for patients under active oncology management within UK NHS. At both sites involved in the study, HCP satisfaction with the pathway was overall high, with expression of readiness for broader rollout with number of benefits identified in previous survey. Our clinician satisfaction data may reflect bias regarding those clinicians electing to participate in the survey; furthermore, the two trusts involved in the study had already established mainstreamed pathways for BC GT and thus their clinician population may not be reflective nationally. Improvements in HCP feedback relating to turnaround times were observed compared to the pilot study, likely through improvements in laboratory test turnaround times (which improved over the study period following COVID19-related laboratory delays and optimisation of copy number variant analysis for saliva-derived DNA).

In terms of the patient population, we offered GT to all women attending for BC diagnosis, management, or follow-up. Under current UK guidelines, fewer than 20% of women with BC are currently eligible for BRCA-testing (22). UK health economic analysis undertaken in 2018 suggest universal testing of BC patients to be economically impactful within NICE willingness to pay thresholds of £30,000/QALY up to a per-patient cost of £1,626 (payer perspective) and £1,868 (societal perspective) (23). Of note, recent guidance from the American Society of Clinical Oncology (ASCO) has recommended testing for *BRCA1* and *BRCA2* in all BC cases up to the age of 65, including those diagnosed historically(24). However, detection of PVs will be higher in cases with younger onset, higher-grade, bilateral and/or hormone-receptor negative disease, and where a relevant family history is present (22). There is an inherent tension in balancing enhancement of detection rate against the complexity and imperfect sensitivity incurred from test eligibility criteria.

Nevertheless, any substantial expansion in BRCA-testing of BC cases, for example testing of all cases arising age ≤65 years, would require higher-throughput clinical and laboratory systems. We propose that such systems should (i) incorporate more generic patient information materials and clinical workflows to enable scale, (ii) retain availability where required of expert clinical input individualised to patient requirements and (iii) attune flexibly to local clinical pathways and informatic workflows, especially as GT timed at point of diagnosis requires efficient timely delivery. Additional work is required to understand necessary facilitators and adaptations to optimise equity of access; pathway-specific health economic analyses will also be important.

Overall, this study demonstrates that, supported by a genetic counselling hotline, a fully-digital pathway may be a suitable alternative to conventional models of pre-test information-giving, sampling and consenting, and could support end-to-end management of genetic testing for a large proportion of BC patients. Where expansion of germline genetic testing is limited by clinical capacity, pathways such as BRCA-DIRECT, implemented within mainstream oncology clinics may offer a viable and acceptable approach for the majority of patients to minimise the patient-facing and administrative burdens to clinicians of BRCA-testing, whilst providing flexible patient access to clinical genetics expertise. Thus enabling both clinical genetics and oncology professional time to be utilised more effectively for management of those with a positive result or supporting a minority of patients for whom a standardised, digital approach is unsuitable.

## ETHICS

This study was approved for sponsorship by The Institute of Cancer Research/Royal Marsden NHS Foundation Trust Joint Committee for Clinical Research. The study received favourable opinion from the London—Chelsea Research Ethics Committee (REC) on 4 January 2021 (REC reference: 20/LO/1200) and full Health Research Authority (HRA) and Health and Care Research Wales (HCRW) approval on 4 January 2021. Participants gave informed consent to participate in the study before taking part.

## DATA AVAILABILITY

The datasets generated and/or analysed during the current study are available from the corresponding author on reasonable request.

## ACKNOWLEDGEMENTS

The authors would like to thank: The members of the patient and public involvement group for their input to the design and delivery of the study. The patients, research and clinical teams at the Royal Marsden NHS Foundation Trust and Manchester University NHS Foundation Trust for their involvement in recruiting and implementing the study pathway. The team at the Centre for Molecular Pathology (Institute of Cancer Research/Royal Marsden NHS Foundation Trust) for establishing and validating the saliva-based germline genetic testing pathway, enabling accredited NHS testing to be performed within this study. The Trial Steering Committee for their scientific oversight and monitoring of the study and outputs.

## REFERENCES

1. Antoniou AC, Foulkes WD, Tischkowitz M. Breast-cancer risk in families with mutations in PALB2. N Engl J Med. 2014;371(17):1651–2.

2. Antoniou AC, Foulkes WD, Tischkowitz M. Breast cancer risk in women with PALB2 mutations in different populations. The Lancet Oncology. 2015;16(8):e375–6.

3. Kuchenbaecker KB, Hopper JL, Barnes DR, Phillips KA, Mooij TM, Roos-Blom MJ, et al. Risks of Breast, Ovarian, and Contralateral Breast Cancer for BRCA1 and BRCA2 Mutation Carriers. JAMA. 2017;317(23):2402–16.

4. England N. National Genomic Test Directory: Testing Criteria for Rare and Inherited Disease. Online2023.

5. Heather JM, Chain B. The sequence of sequencers: The history of sequencing DNA. Genomics. 2016;107(1):1–8.

6. Tung NM, Boughey JC, Pierce LJ, Robson ME, Bedrosian I, Dietz JR, et al. Management of Hereditary Breast Cancer: American Society of Clinical Oncology, American Society for Radiation Oncology, and Society of Surgical Oncology Guideline. Journal of Clinical Oncology. 2020;38(18):2080–106.

7. Cortesi L, Rugo HS, Jackisch C. An Overview of PARP Inhibitors for the Treatment of Breast Cancer. Targeted Oncology. 2021;16(3):255–82.

8. Excellence NIfHaC. Olaparib for adjuvant treatment of BRCA mutation-positive HER2-negative high-risk early breast cancer after chemotherapy Online 2023 10 May 2023 Contract No.: [TA886].

9. Tutt ANJ, Garber JE, Kaufman B, Viale G, Fumagalli D, Rastogi P, et al. Adjuvant Olaparib for Patients with BRCA1- or BRCA2-Mutated Breast Cancer. New England Journal of Medicine. 2021;384(25):2394–405.

10. NICE. Olaparib for adjuvant treatment of BRCA mutation-positive HER2-negative high-risk early breast cancer after chemotherapy. In: guidance Ta, editor. 2023.

11. NICE. Talazoparib for treating HER2-negative advanced breast cancer with germline BRCA mutations. In: Guidance TA, editor. 2024.

12. Slade I, Riddell D, Turnbull C, Hanson H, Rahman N. Development of cancer genetic services in the UK: A national consultation. Genome Med. 2015;7(1):18.

13. Hallowell N, Wright S, Stirling D, Gourley C, Young O, Porteous M. Moving into the mainstream: healthcare professionals’ views of implementing treatment focussed genetic testing in breast cancer care. Familial cancer. 2019.

14. Torr B, Jones C, Choi S, Allen S, Kavanaugh G, Hamill M, et al. A digital pathway for genetic testing in UK NHS patients with cancer: BRCA-DIRECT randomised study internal pilot. Journal of Medical Genetics. 2022;59(12):1179–88.

15. Sealed Envelope Ltd. Create a blocked randomisation list. https://www.sealedenvelope.com/simple-randomiser/v1/lists 2021 [

16. Speilberger C, Gorsuch R, Lushene R, Vagg P, Jacobs G. Manual for the Stait-Trait Anxiety Inventory (Form Y1-Y2). Palo Alto, CA: . Consulting Psychology Press. 1983.

17. Carleton RN, Norton MA, Asmundson GJ. Fearing the unknown: a short version of the Intolerance of Uncertainty Scale. Journal of anxiety disorders. 2007;21(1):105–17.

18. Lee W, Shickh S, Assamad D, Luca S, Clausen M, Somerville C, et al. Patient-facing digital tools for delivering genetic services: a systematic review. Journal of Medical Genetics. 2023;60(1):1–10.

19. Manchanda R, Burnell M, Loggenberg K, Desai R, Wardle J, Sanderson SC, et al. Cluster-randomised non-inferiority trial comparing DVD-assisted and traditional genetic counselling in systematic population testing for BRCA1/2 mutations. J Med Genet. 2016;53(7):472–80.

20. Swisher EM, Rayes N, Bowen D, Peterson CB, Norquist BM, Coffin T, et al. Remotely Delivered Cancer Genetic Testing in the Making Genetic Testing Accessible (MAGENTA) Trial: A Randomized Clinical Trial. JAMA Oncology. 2023;9(11):1547–55.

21. Sie AS, van Zelst-Stams WA, Spruijt L, Mensenkamp AR, Ligtenberg MJ, Brunner HG, et al. More breast cancer patients prefer BRCA-mutation testing without prior face-to-face genetic counseling. Familial cancer. 2014;13(2):143–51.

22. Evans DG, Woodward ER, Burghel GJ, Allen S, Torr B, Hamill M, et al. Population-based germline testing of BRCA1, BRCA2, and PALB2 in breast cancer patients in the United Kingdom: Evidence to support extended testing, and definition of groups who may not require testing. Genetics in Medicine Open. 2024;2:100849.

23. Sun L, Brentnall A, Patel S, Buist DSM, Bowles EJA, Evans DGR, et al. A Cost-effectiveness Analysis of Multigene Testing for All Patients With Breast Cancer. JAMA Oncology. 2019;5(12):1718–30.

24. Bedrosian I, Somerfield MR, Achatz MI, Boughey JC, Curigliano G, Friedman S, et al. Germline Testing in Patients With Breast Cancer: ASCO–Society of Surgical Oncology Guideline. Journal of Clinical Oncology. 2024;42(5):584–604.

