## Supplementary material for "BRCA-DIRECT digital pathway for diagnostic germline genetic testing within a UK breast oncology setting: a randomised, non-inferiority trial"

**Title:**

**Affiliations**

### Supplementary Figure 1: Adjusted differences between the fully-digital arm compared to the partially-digital arm.

δ Indicates the non-inferiority margin.

#### (a) uptake of genetic testing

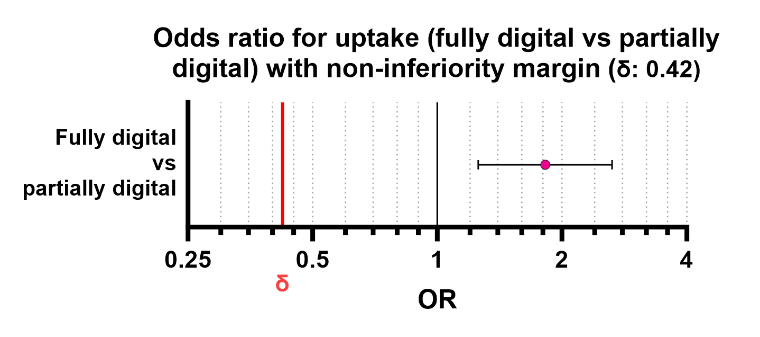

OR = Odds ratio

(b) mean knowledge score
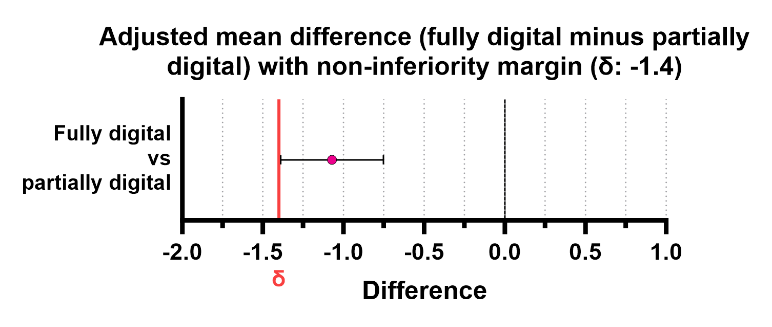

#### (c) mean anxiety scores

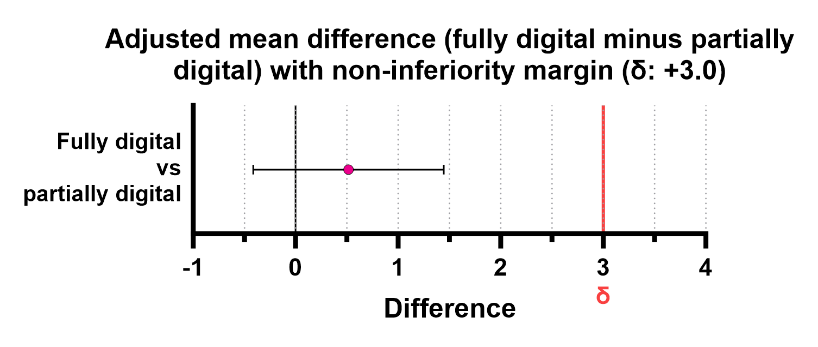

(d) mean satisfaction scores
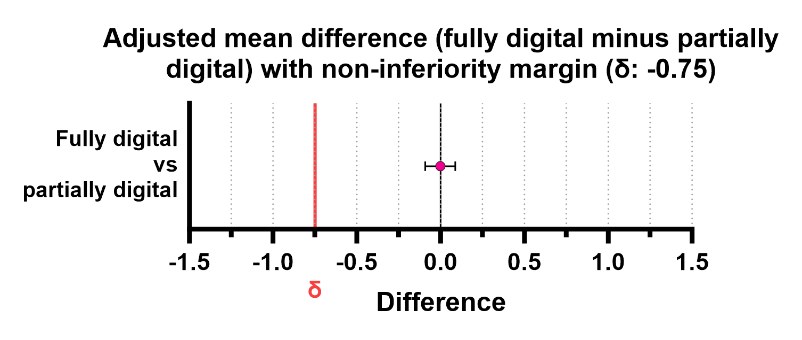

T1 = 1-day post genetic test consent

T2 = 7-days post genetic test results

T3 = 28-days post genetic test results

Supplementary Figure 2: Baseline and adjusted mean knowledge scores in the fully- and partially-digital arms over time

|  | Partially | Fully digital |
| --- | --- | --- |
| Baseline | 5.14677 | 5.4718 |
| T1 | 9.35693 | 8.28589 |
| T3 | 9.05126 | 7.98023 |

Supplementary **Figure 3:** Baseline and adjusted mean anxiety scores in the fully- and partially-digital arms over time

|  | Partially | Fully digital |
| --- | --- | --- |
| Baseline | 41.7513 | 41.9964 |
| T1 | 38.8483 | 39.3633 |
| T2 | 37.4066 | 37.9216 |
| T3 | 36.9858 | 37.5008 |

### Supplementary Table 1: Methods of evaluating outcomes for assessing non-inferiority of digital information with the timepoints of assessment.

| **Outcome** | **Measure** | **Timepoint of assessment** | | | | **Non-inferiority Margin** (sample size required for 80% power) |
| --- | --- | --- | --- | --- | --- | --- |
|  |  | Baseline | 1-day after genetic test consent (T1) | 7-days after receiving the genetic test result (T2) | 28-days after receiving the genetic test result (T3) |  |
| **Genetic test uptake** | Proportion of participants consenting to genetic testing | N/A | | | | -5.5%  (934) |
| **Anxiety** | State Trait Anxiety Inventory  20-item validated survey, producing state and trait scores between 20 and 80, with higher scores reflecting greater anxiety. ^19^ | - Trait and state anxiety | - State anxiety | - State anxiety | - State anxiety | +3  (636) |
|  | Intolerance of uncertainty  ^20^ |  |  |  |  |  |
| **Knowledge** | 14-item study specific questionnaire, with true or false answers. Participants could also answer ‘don’t know’. Scores reflected total correct answers out of a possible 14. |  |  |  |  | -1.4  (56) |
| **Participant satisfaction** | 10-item study specific survey.  Focussed question on satisfaction with the method of receiving pre-test information Scored on a 5-point Likert scale (1- very unsatisfied; 5 – very satisfied). |  |  | (invited at T2 with reminder at T3 if incomplete) | | -0.75  (38) |

### Supplementary Table 2: Non-inferiority margin assumptions and power

|  | **Attrition** | **Predicted available participants (following attrition)** | **Assumptions**  **(reference *)** | **Power** | **Sample size required for 80% power (digital: clinician = 1:1)** | **Significance level (one-sided)** | **Non-inferiority margin** | **Notes** |
| --- | --- | --- | --- | --- | --- | --- | --- | --- |
| **Test uptake** | 0% | 1000 | 90% test uptake in both groups | 80-85% with 1000 participants | 892 | 0.05 | 5% | Calculated using [http://www.hwasoon.kim/NISSC/#!/binary](http://www.hwasoon.kim/NISSC/) |
| **STAI state T1** | 0% | 1000 | Mean 43.2, SD 13.5 ^1^  *(range 0 – 100)* | >95% with 1000 participants | 500 | 0.05 | 3 | Calculated using [http://www.hwasoon.kim/NISSC/#!/continuous](http://www.hwasoon.kim/NISSC/) |
| **STAI state T2** | 10% | 900 | Mean 43.2, SD 13.5 ^1^  (range 0 – 100) | >95% with 900 participants | 500 | 0.05 | 3 |  |
| **STAI state T3** | 20% | 800 | Mean 43.2, SD 13.5 ^1^  (range 0 – 100) | 90-95% with 800 participants | 500 | 0.05 | 3 |  |
| **Patient satisfaction** | 20% | 800 | Mean 3.75, SD 1  (out of 5 – Likert scale)  ^2^ | >95% with 800 participants | 44 | 0.05 | 0.75 |  |
| **Knowledge score T1** | 20% | 800 | Mean 5.71, SD 1.55  (out of a possible 10) ^3^ | >95% with 800 participants | 30 | 0.05 | 1.40 |  |

### Supplementary Table 3: Sensitivity and subgroup analyses

#### (a) Genetic test uptake

|  | | Category representation (all participants) | | subgroup#intervention interaction | | | | Intervention effect in subgroup | | |
| --- | --- | --- | --- | --- | --- | --- | --- | --- | --- | --- |
|  |  | n | % | estimate | 95% CI | p-value | model n | estimate | 95% CI | p-value |
| Reported breast cancer history in first degree relatives | No | 789 | 74.57 | Reference category | | | | *No effect observed* | | |
|  | Yes | 269 | 25.43 | 0.62 | 0.18 to 2.18 | 0.455 | 1058 |  |  |  |
|  | Total | 1058 | 100.00 |  | | | |  |  |  |
| Reported breast cancer history in second degree relatives | No | 707 | 66.82 | Reference category | | | | *No effect observed* | | |
|  | Yes | 351 | 33.18 | 0.34 | 0.09 to 1.34 | 0.124 | 1058 |  |  |  |
|  | Total | 1058 | 100.00 |  | | | |  |  |  |
| Reported other cancer history in relatives | No | 886 | 83.74 | Reference category | | | | *No effect observed* | | |
|  | Yes | 172 | 16.26 | 0.47 | 0.07 to 3.22 | 0.445 | 1058 |  |  |  |
|  | Total | 1058 | 100.00 |  | | | |  |  |  |
| Breast cancer status | Newly diagnosed | 667 | 58.61 | Reference category | | | | *No effect observed* | | |
|  | In follow up | 400 | 35.15 | 1.41 | 0.64 to 3.07 | 0.392 | 1138 |  |  |  |
|  | Metastatic | 71 | 6.24 | 0.44 | 0.10 to 1.98 | 0.286 | 1138 |  |  |  |
|  | Total | 1138 | 100.00 |  | | | |  |  |  |
| Genetic test result | Negative | 969 | 97.00 | Not modelled | | | | | | |
|  | Pathogenic | 30 | 3.00 |  |  |  |  |  |  |  |
|  | Total | 999 | 100.00 |  |  |  |  |  |  |  |
| Method of receiving result from genetic test | Digitally | 942 | 94.29 | Not modelled | | | | | | |
|  | Telephone appointment | 57 | 5.71 |  |  |  |  |  |  |  |
|  | Total | 999 | 100.00 |  |  |  |  |  |  |  |

#### (b) Knowledge

|  | | | Category representation (all participants) | | | subgroup#intervention interaction | | | | Intervention effect in subgroup | | |
| --- | --- | --- | --- | --- | --- | --- | --- | --- | --- | --- | --- | --- |
|  |  |  | n | | % | estimate | 95% CI | p-value | model n | estimate | 95% CI | p-value |
| Reported breast cancer history in first degree relatives | No | 789 | | 74.57 | | Reference category | | | | *No effect observed* | | |
|  | Yes | 269 | | 25.43 | | -0.45 | -1.17 to 0.28 | 0.225 | 989 |  | | |
|  | Total | 1058 | | 100.00 | |  | | | |  | | |
| Reported breast cancer history in second degree relatives | No | 707 | | 66.82 | | Reference category | | | | *No effect observed* | | |
|  | Yes | 351 | | 33.18 | | 0.23 | -0.44 to 0.90 | 0.496 | 989 |  | | |
|  | Total | 1058 | | 100.00 | |  | | | |  | | |
| Reported other cancer history in relatives | No | 886 | | 83.74 | | Reference category | | | | *No effect observed* | | |
|  | Yes | 172 | | 16.26 | | 0.74 | -0.12 to 1.59 | 0.093 | 989 |  | | |
|  | Total | 1058 | | 100.00 | |  | | | |  | | |
| Breast cancer status | Newly diagnosed | 667 | | 58.61 | | Reference category | | | | *No effect observed* | | |
|  | In follow up | 400 | | 35.15 | | 0.30 | -0.38 to 0.98 | 0.382 | 989 |  | | |
|  | Metastatic | 71 | | 6.24 | | 0.45 | -0.92 to 1.82 | 0.520 |  |  |  |  |
|  | Total | 1138 | | 100.00 | |  | | | |  | | |
| Test result | Negative | 969 | | 97.00 | | Reference category | | | | *No effect observed* | | |
|  | Pathogenic | 30 | | 3.00 | | 1.20 | -0.64 to 3.05 | 0.202 | 987 |  | | |
|  | Total | 999 | | 100.00 | |  | | | |  | | |
| Method of receiving result | Digitally | 942 | | 94.29 | | Reference category | | | | *No effect observed* | | |
|  | Telephone appointment | 57 | | 5.71 | | 0.77 | -0.58 to 2.12 | 0.266 | 987 |  | | |
|  | Total | 999 | | 100.00 | |  | | | |  | | |

#### (c) Anxiety

|  | | Category representation (all participants) | | | subgroup#intervention interaction | | | | Intervention effect in subgroup | | |
| --- | --- | --- | --- | --- | --- | --- | --- | --- | --- | --- | --- |
|  |  | n | % | estimate | | 95% CI | p-value | model n | estimate | 95% CI | p-value |
| Reported breast cancer history in first degree relatives | No | 789 | 74.57 | Reference category | | | | | *No effect observed* | | |
|  | Yes | 269 | 25.43 | 0.13 | | -2.00 to 2.25 | 0.908 | 995 |  | | |
|  | Total | 1058 | 100.00 |  | | | | |  | | |
| Reported breast cancer history in second degree relatives | No | 707 | 66.82 | Reference category | | | | | *No effect observed* | | |
|  | Yes | 351 | 33.18 | -0.09 | | -2.05 to 1.87 | 0.929 | 995 |  | | |
|  | Total | 1058 | 100.00 |  | | | | |  | | |
| Reported other cancer history in relatives | No | 886 | 83.74 | Reference category | | | | | *No effect observed* | | |
|  | Yes | 172 | 16.26 | -0.34 | | -2.83 to 2.15 | 0.79 | 995 |  | | |
|  | Total | 1058 | 100.00 |  | | | | |  | | |
| Breast cancer status | Newly diagnosed | 667 | 58.61 | Reference category | | | | | *No effect observed* | | |
|  | In follow up | 400 | 35.15 | -2.10 | | -4.07 to -0.12 | 0.038 | 995 |  | | |
|  | Metastatic | 71 | 6.24 | -1.73 | | -5.74 to 2.28 | 0.399 |  |  |  |  |
|  | Total | 1138 | 100.00 |  | | | | |  | | |
| Test result | Negative | 969 | 97.00 | Reference category | | | | | *No effect observed* | | |
|  | Pathogenic | 30 | 3.00 | -1.06 | | -6.43 to 4.30 | 0.698 | 993 |  | | |
|  | Total | 999 | 100.00 |  | | | | |  | | |
| Method of receiving result | Digitally | 942 | 94.29 | Reference category | | | | | *No effect observed* | | |
|  | Telephone appointment | 57 | 5.71 | -0.56 | | -4.51 to 3.38 | 0.779 | 993 |  | | |
|  | Total | 999 | 100.00 |  | | | | |  | | |

#### (d) Satisfaction

|  | | Category representation (all participants) | | subgroup#intervention interaction | | | | Intervention effect in subgroup | | |
| --- | --- | --- | --- | --- | --- | --- | --- | --- | --- | --- |
|  |  | n | % | estimate | 95% CI | p-value | model n | estimate | 95% CI | p-value |
| Reported breast cancer history in first degree relatives | No | 789 | 74.6 | Reference category | | | | *No effect observed* | | |
|  | Yes | 269 | 25.4 | -0.14 | -o.34 to 0.07 | 0.198 | 908 |  | | |
|  | Total | 1058 | 100.0 |  | | | |  | | |
| Reported breast cancer history in second degree relatives | No | 707 | 66.8 | Reference category | | | | *No effect observed* | | |
|  | Yes | 351 | 33.2 | 0.06 | -0.12 to 0.24 | 0.511 | 908 |  | | |
|  | Total | 1058 | 100.0 |  | | | |  | | |
| Reported other cancer history in relatives | No | 886 | 83.7 | Reference category | | | | *No effect observed* | | |
|  | Yes | 172 | 16.3 | 0.10 | -0.15 to 0.34 | 0.441 | 908 |  | | |
|  | Total | 1058 | 100.0 |  | | | |  | | |
| Breast cancer status | Newly diagnosed | 667 | 58.6 | Reference category | | | | *No effect observed* | | |
|  | In follow up | 400 | 35.2 | 0.10 | -0.10 to 0.29 | 0.330 | 908 |  | | |
|  | Metastatic | 71 | 6.2 | -0.02 | -0.43 to 0.38 | 0.909 | 908 |  |  |  |
|  | Total | 1138 | 100.0 |  | | | |  | | |
| Test result | Negative | 969 | 97.0 | Reference category | | | | *No effect observed* | | |
|  | Pathogenic | 30 | 3.0 | -0.28 | -0.80 to 0.24 | 0.292 | 908 |  | | |
|  | Total | 999 | 100.0 |  | | | |  | | |
| Method of receiving result | Digitally | 942 | 94.3 | Reference category | | | | *No effect observed* | | |
|  | Telephone appointment | 57 | 5.7 | -0.21 | -0.59 to 0.17 | 0.276 | 908 |  | | |
|  | Total | 999 | 100.0 |  | | | |  | | |

### References

1	Mansel RE, Fallowfield L, Kissin M et al. Randomized multicenter trial of sentinel node biopsy versus standard axillary treatment in operable breast cancer: the ALMANAC Trial. J Natl Cancer Inst 2006; 98 (9): 599-609.

2	Ware JE, Jr., Hays RD. Methods for measuring patient satisfaction with specific medical encounters. Medical care 1988; 26 (4): 393-402.

3	Meisel SF, Freeman M, Waller J et al. Impact of a decision aid about stratified ovarian cancer risk-management on women’s knowledge and intentions: a randomised online experimental survey study. BMC public health 2017; 17 (1): 882.
